# Shared genetic and molecular architecture between insulin resistance and cognitive performance

**DOI:** 10.64898/2026.07.15.26358124

**Authors:** Alfonso Martone, Nina Roth Mota, Barbara Šakić, Marieke Klein, Barbara Franke, Giuseppe Fanelli, Janita Bralten

## Abstract

Insulin signalling contributes to neurodevelopment and brain function, and insulin resistance (IR)-related traits are associated with cognitive performance. However, the genetic architecture shared across specific cognitive domains and IR-related phenotypes remains insufficiently defined. We analysed large-scale genome-wide association study summary statistics for 11 IR-related traits (N=53,334–933,970) and 10 cognitive measures (N=28,156–436,853) to quantify global and local genetic correlations, fine-map shared association signals, and annotate implicated genes and drug–gene interactions. Pairwise global and local genetic correlations were estimated, and shared high-confidence variants were prioritised using the multivariate Sum of Single Effects model. Positional and expression quantitative trait locus mapping was performed, and implicated genes were examined through functional annotation, tissue enrichment, and drug–gene interaction analyses. Low-to-moderate genetic correlations were observed between six IR-related traits and seven cognitive measures (|r_g_|=0.08-0.34), with predominantly opposite directions, except for correlations involving visual declarative short-term memory. Local genetic correlations showed mixed effect directions across most trait pairs, and multivariate fine-mapping prioritised 696 shared likely causal variants with high posterior support. Gene annotation indicated enrichment in several pathways, including immune-related, signal transduction, neurogenesis, neurotransmitter metabolism, receptor regulation, and lipid and cholesterol metabolism regulation. Implicated genes were expressed across various brain regions and showed prior associations with neuropsychiatric and cardiometabolic conditions. Several drug-gene interactions were identified, involving immunomodulatory and anti-inflammatory compounds. These findings indicate widespread heterogeneous genetic overlap between IR-related traits, particularly body mass index and waist-to-hip ratio, and cognitive measures of general intelligence, processing speed, and short-term visual declarative memory. The findings prioritise apolipoprotein-related lipid transport and inflammatory and oxidative stress pathways as candidate mechanisms linking cognitive, cardiometabolic, and neuropsychiatric phenotypes.

## Introduction

A growing body of epidemiological, clinical, and experimental evidence supports an association between insulin resistance (IR) and cognitive performance. IR denotes a reduced biological response to insulin in target tissues and can be reflected by compensatory hyperinsulinaemia, altered glucose homeostasis, or related metabolic phenotypes (1). IR-related conditions, like type 2 diabetes mellitus (T2DM), metabolic syndrome (MetS), and obesity, are consistently associated with poorer performance in several cognitive domains, including global cognitive functioning, verbal and numeric reasoning, executive functions, working memory, processing speed. auditory attention, and word comprehension (2–7). T2DM and cardiovascular diseases were also shown to predict processing speed performance in a machine-learning study of a population-based cohort (8). Consistently, worse cognitive performance in different domains has also been associated with somatic features of IR, including measures of body adiposity (*e.g.* body-mass index (BMI) and waist-to-hip ratio (WHR)), blood glucose and insulin levels, glycated haemoglobin (HbA1c) (9–12). Although not all cognitive domains and tasks show the same relationship with IR (3,9,11) and a better performance at baseline in visual declarative short-term memory has been reported, diabetic subjects show a steeper decline compared to non-diabetic subjects (3).

Since IR as well as cognitive performance are multifactorial phenotypes with a moderate to large genetic component (13–16), understanding the shared genetic architecture of IR and cognition can be particularly informative on shared mechanisms, especially considering that both IR and cognitive alterations are often reported in neuropsychiatric disorders (17,18). Cognitive performance in specific domains varies and overlaps among psychiatric diagnoses (17), and cognitive impairment is frequently detected at the onset of psychiatric disorders or as residual and difficult-to-treat symptoms, often associated with treatment response (17,19–21). Regarding IR, insulin signaling has a central role in key neural networks, and specific structural and functional alterations have been linked to IR in the brain. Hyperglycaemia has been associated with structural abnormalities in the PFC (22) and other brain regions (23), and chronic exposure to elevated glucose levels may lead to an accelerated cognitive decline (24,25). The insulin system in the brain appears to actively modulate information processing, attention shift, task and behavioural flexibility, memory, and processing speed through its effect on the mesostriatal and mesocortical pathways (26,27). IR-associated disruptions in the brain’s dopaminergic system have been linked to motivational deficits and depressive symptoms (28). This is consistent with evidence linking IR to neuropsychiatric symptoms pathophysiology (29,30); IR may also contribute to developmental trajectories connecting prenatal and early-life adversities with later psychopathology, although causal mediation remains to be established (31,32).Thus, variation across specific cognitive domains and the cognitive-metabolic axis may represent transdiagnostic and more proximal intermediate phenotypes relevant to psychiatric disorders, with possible therapeutic and diagnostic potential.

Despite converging phenotypic and experimental evidence of the relevance of IR in cognition and neuropsychiatric disorders, little is known about the genetic basis underlying the association between cognition and IR and which cognitive domains are genetically linked to IR. In fact, previous genetic studies have generally examined individual metabolic traits, single cognitive outcomes, genome-wide average correlations, or candidate molecular pathways (26,33–35). The regional distribution, direction, and variant-level basis of the genetic overlap across a broader range of IR-related and cognitive phenotypes remain insufficiently studied.

Therefore, to investigate which cognitive domains genetically link to IR and to characterize the common genetic background of cognitive performance and IR, we conducted a large-scale analysis of shared genetic signals between 11 IR-related traits and 10 cognitive domains using genome-wide association study (GWAS) summary statistics. We first quantified global genetic correlations to assess pairwise average genome-wide correlation. We then examined local patterns of genetic correlation to identify specific genomic regions contributing to shared liability, including loci with opposing effect directions that may be overshadowed at the global level. Finally, multivariate fine-mapping and functional annotation were applied to prioritise shared likely causal variants, implicated genes, biological pathways, and druggable molecular targets. This multi-step approach was designed to provide a detailed (and domain-specific) characterisation of the genetic and molecular links between IR and cognition.

## Material and Methods

### Input datasets

Publicly available European-ancestry summary statistics of the largest and most recent GWASs for 10 cognitive tasks and 11 IR-related conditions and traits were collected (see Supplementary Table S1). Input datasets were selected based on phenotypic relevance, sample size (N>10,000), and test-retest reliability and correlation with the reference cognitive construct (Pearson’s R ≥0.3) (36,37). To retain traits with sufficient common-variant signal for downstream analyses, we excluded traits with SNP-based heritability (*h^2^*) <0.04.

GWAS summary statistics underwent quality-control, retaining only variants with minor allele frequency ≥0.01 and imputation quality ≥0.9 (where this information was available), and matching alleles listed in the HapMap Project Phase 3. Strand-ambiguous SNPs, indels, and variants missing required fields or with invalid effect estimates, standard errors, or allele frequencies were excluded. Where lower values corresponded to better cognitive performance, e.g. for mean Reaction Time and the trail-making tests A and B, z-score signs were flipped in the original papers for consistency of effect directions across traits (16). For consistency, GWAS z-scores for Stumvoll’s Insulin Sensitivity Index (ISI) were reversed in sign as well, given the inverse relationship between ISI and the other IR-related traits.

### Global genetic correlation

The *ldsc* function from the GenomicSEM package for R (https://github.com/GenomicSEM/) was used to calculate intercepts and bivariate global genetic correlation (r_g_) through Linkage Disequilibrium SCore regression (LDSC) between each of the 10 cognitive traits and each of the 11 IR-related traits (38). Pre-computed linkage disequilibrium (LD) scores from the European subset of the 1,000 Genomes Project were used for the regression weights (39,40). Conservative Bonferroni multiple testing correction was applied, correcting for the number of performed tests (α=0.05/(10*11)=0.00045).

### Local genetic correlation

The LAVA (Local Analysis of [co]Variant Association; https://github.com/josefin-werme/LAVA) R package was utilised to assess the pairwise patterns of local genetic correlation between the cognitive and IR-related traits (41).

Following the approach reported by the authors (https://github.com/cadeleeuw/lava-partitioning) (42), 2495 semi-independent genomic regions of ∼1Mb in minimal LD were generated using the European subset of the 1000 Genomes Project, phase 3 (40) with a MAF threshold of 0.01. For each genomic region, *h^2^_SNP_* was evaluated through univariate analyses, and genomic regions with univariate p-value < 10^-4^ were selected for subsequent bivariate analyses (41). Pairwise local genetic correlations (ρ) were then estimated between the cognitive and IR-related traits at each of the selected genomic regions. Intercepts from the LDSC analyses were provided to allow LAVA to account for sample overlap, as these represent an estimate of sampling correlation between datasets (39). Finally, LAVA bivariate analyses results were corrected for multiple testing controlling for the expected proportion of false positives through False Discovery Rate (FDR) < 0.05 considering all bivariate tests performed. This was done in line with previous publications (29,43) and in consideration of the multi-step approach applied here which adopts stringent follow-up analyses.

### Multivariate fine-mapping of shared likely causal variants

To prioritise shared likely causal variants within regions showing significant local genetic correlation, the multi-trait fine-mapping R package *mvsusieR* was used (44) (https://github.com/stephenslab/mvsusieR). The multivariate Sum of Single Effects (mvSuSiE) tool represents a multivariate extension of the SuSiE model (45), combining the Multivariate Adaptive Shrinkage (mash) statistical approach (46) to allow the model to flexibly learn the patterns of shared genetic effects across different traits (44).

In accordance with mvSuSiE documentation (https://stephenslab.github.io/mvsusieR/), a set of eight matrices corresponding to prior knowledges was provided, including an identity matrix (which assumes that all the traits are independent), the LDSC covariance matrix (the genetic relationship observed between the traits), and six covariance matrices derived from the trait-specific SNP z-scores as suggested by the developers (46). A final weighted mixture prior was calculated using *mash* (46) assuming an equal weight for each of the input matrices, thus allowing mvSuSiE to estimate the best-fitting model. The analyses were then performed looping through all loci in the areas that showed significant correlations in the LAVA bivariate analyses. Each significant locus was extracted with the corresponding traits that were correlated through it, running mvSuSiE on each of them by providing, at the same time, the locus z-score matrix (a matrix with the GWAS-derived z-scores for each SNP in the locus), the LD matrix of the locus, the smallest GWAS sample size among the included traits, and the weighted mixture prior. The maximum number of single-effect components was set to 10, with a convergence tolerance of 0.001 and a maximum of 1000 iterations.

The resulting credible sets were collected and screened as described elsewhere (44). Each credible set represents an independent association signal in the data, designed to capture at least one putative causal SNP with high probability for the observed association. SNPs with a cross-trait posterior inclusion probability (PIP) above 0.95 were deemed as likely causal for at least one trait, whereas a local false sign rate (lfsr) <0.01 was used to identify which traits are affected by each credible set. In case the credible set contained more than one SNP, the average lfsr was used (44).

### Positional gene mapping

The identified credible sets were positionally mapped to the corresponding genes through the web-based variant annotation tool SNPnexus (47,48). The annotation query included positional mapping (within a genomic window of ±10kb), Ensembl gene and protein consequences, SIFT-based evaluation of non-synonymous coding SNPs effect on protein function, Genetic Association Database (GAD), NHGRI Catalogue of Published GWAS for phenotype and disease association, and Reactome database for pathway analysis (https://www.snp-nexus.org/v4/).

### eQTL-based gene mapping

For the eQTL-based gene mapping, the *loci2path* R package (v.1.3.1, https://github.com/StanleyXu/loci2path) was used (49) to map genes regulated by eQTLs located within genomic regions showing FDR-significant bivariate local genetic correlations in LAVA across 13 areas of the central nervous system (*i.e*. amygdala, anterior cingulate cortex, caudate nucleus, cerebellar hemisphere, cerebellum, brain cortex, frontal cortex Brodmann Area (BA) 9, hippocampus, hypothalamus, nucleus accumbens, putamen, spinal cord, and substantia nigra). The eQTL data for these areas were obtained from the Genotype-Tissue Expression (GTEx) project (GTEx v10, GRCh38/hg38) (https://gtexportal.org/home/downloads/adult-gtex/qtl), lifting the eQTL coordinates to the GRCh37/hg19 genomic build using the UCSC LiftOver tool (https://genome-store.ucsc.edu) to match the genomic build of the used summary statistics. We used GTEx v10 significant cis-eQTL variant–gene pairs for each tissue, as defined by the GTEx tissue-specific false discovery rate procedure.

### Functional annotation and drug targetability

Functional Mapping and Annotation (FUMA) GENE2FUNC module (https://fuma.ctglab.nl/gene2func)(50) was used to assess tissue-expression and gene-set enrichment of both positional and eQTL mapped genes (both separately and combined). All human genes were used as background, and the MHC region was not excluded from analysis. Default parameters were applied, except for the following: Ensembl version v110; GTEx v8 and BrainSpan tissue expression datasets; Benjamini-Hochberg FDR correction for multiple testing.

The list of mapped genes was assessed for drug targetability through the Drug-Gene Interaction Database (DGIdb, https://dgidb.org/) using default settings by taking into account the corresponding interaction score (51). DGIdb interaction scores were used to rank previously reported drug–gene relationships and were not interpreted as measures of therapeutic efficacy or direction of treatment effect.

## Results

### Input datasets

GWAS summary statistics of the cognitive traits were obtained from Wootton et al. 2024 (16), based on data from the large-scale population-based UK Biobank sample. Ten out of the 12 cognitive traits analysed were included; Reaction Time Variability was excluded based on SNP-based heritability (h^2^ =0.03), and Prospective Memory was excluded due to low test reliability (Pearson’s R = 0.224) (36). The included traits cover five general cognitive domains: executive functions, memory, attention, fluid intelligence, and processing speed. The sample size ranged from 28,156 (trail-making test B) to 436,853 (pairs matching test), with a h^2^ = 0.04 - 0.22.

For the IR-related conditions and traits, 11 GWASs were selected, including: obesity, T2DM, MetS, the ISI, Insulin Fold Change (IFC), Fasting Plasma Insulin (FPI), Fasting Plasma Glucose (FPG), glycated haemoglobin levels (HbA1c), 2h glycaemia after an oral glucose test (2hGlu), BMI, and WHR. The total sample sizes of the included GWASs varied between 63,396 (2hGlu) and 933,970 (T2DM, 853,816 controls), with an h^2^ = 0.06 - 0.27.

A full list of the included summary statistics with additional information can be found in Supplementary Table S1; for more details about the specifics of the cognitive tasks or concerning the IR-related traits summary statistics, see the original publications (16,50,52–56).

### Genome-wide genetic sharing between insulin resistance and cognitive traits

After Bonferroni correction, 21 pairwise global genetic correlations (r_g_) between the cognitive tasks and IR-related traits were statistically significant, involving fluid intelligence (FI), numeric memory (NM), pairs matching test (PM), matrix pattern recognition (MPR), paired associated learning (PAL), trail-making test B (TMT-B), and symbol digit substitution (SDS). Most showed negative genetic associations with the IR-related traits, going from r_g_=-0.12 (FI-MetS, p=5.94×10^-7^) to r_g_=-0.34 (PAL-ISI, p=5.30×10^-6^). The exception was PM, which was positively correlated with BMI (r_g_=0.14, p=9.96×10^-12^), WHR (r_g_=0.08, p=1.65×10^-4^), and MetS (r_g_=0.13, p=4.83×10^-6^).

For the IR-related phenotypes, correlations involving BMI, WHR, ISI, MetS, Obesity, and T2DM survived Bonferroni correction; WHR showed the highest number of significant associations, genetically correlated to all seven cognitive tests mentioned above, followed by BMI (four significant associations), MetS and T2D (three associations), and ISI and Obesity (two associations).

No significant associations were found for the tower rearranging test (TRT), TMT-A, and reversed mean reaction time (meanRT). All LDSC results are summarised in Table 1.

**Table 1.**
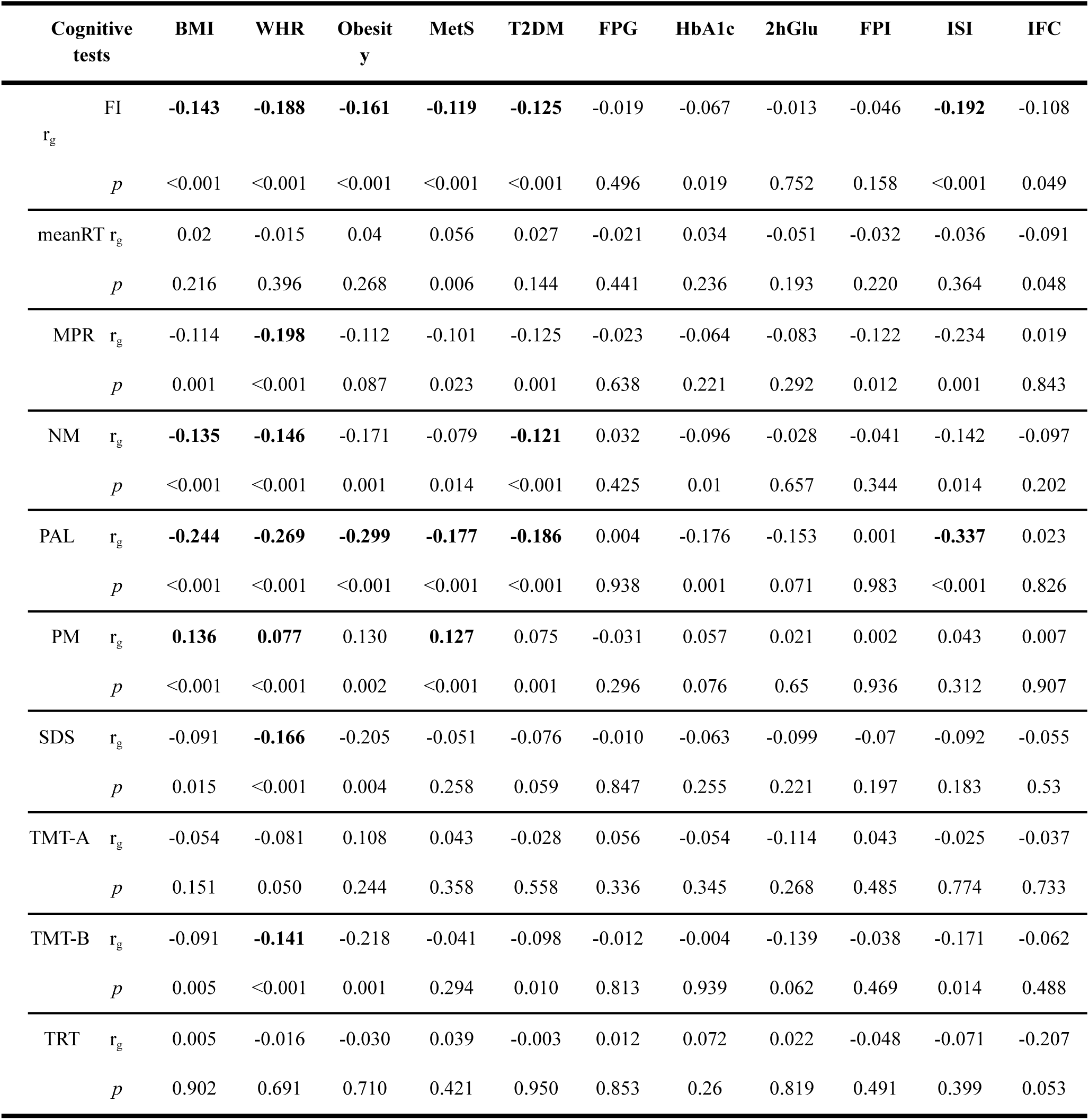
Summary of LDSC results. 2hGlu, 2h glycaemia after an oral glucose challenge; BMI, body mass index; FI, fluid intelligence; FPG, fasting plasma glucose; FPI, fasting plasma insulin; HbA1c, glycated haemoglobin; IFC, insulin fold change; ISI, Stumvoll insulin sensitivity index; IR, insulin resistance; MPR, matrix pattern recognition; meanRT, reaction time (snap game); MetS, metabolic syndrome; NM, numeric memory; PM, pairs matching test; PAL, paired associated learning test; SDS, symbol digit substitution; TMT, trail-making test (A and B); TRT, tower rearranging; T2DM, type 2 diabetes mellitus; WHR, waist-to-hip ratio. r_g_, global genetic correlation, *p*, p-value. Statistically significant results are in **bold** (Bonferroni α=0.00045).

### Local patterns of shared genetic effects

The 311 FDR-significant (q_FDR_<0.05) local bivariate tests exhibited moderate to high degrees of local genetic correlation (|ρ|=0.25-0.98), for a total of 221 unique loci across 63 trait pairs, including also 47 pairs that had not shown a significant global genetic correlation. Five LDSC-significant pairs did not show any local pattern of genetic correlation, namely: Obesity-FI, ISI-PAL, MetS-PAL, Obesity-PAL, and T2DM-PAL. Thirty-six out of the 63 trait pairs registered local genetic correlation in at least two loci. An overview of local genetic correlations is provided in Figure 1 and 3.

**Figure 1.**
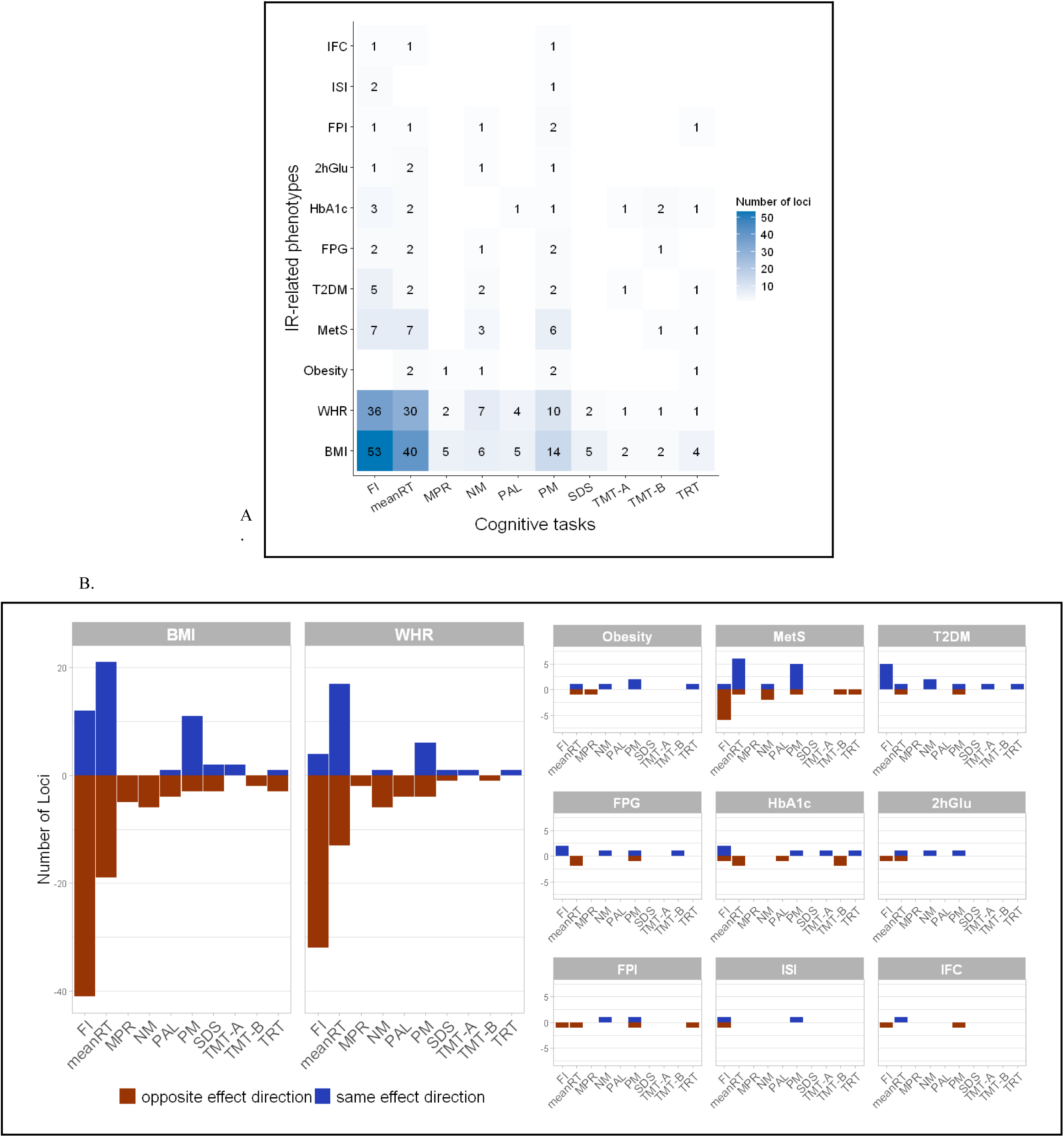
A. Correlation plot showing number of significant shared loci identified at LAVA bivariate analysis. **B.** Barplots showing the number of LAVA-significant loci per trait pairs, divided effect direction. 2hGlu, 2h glycaemia after an oral glucose test; BMI, body mass index; FI, fluid intelligence; FPG, fasting plasma glucose; FPI, fasting plasma insulin; HbA1c, glycated haemoglobin; IFC, insulin fold change; ISI, Stumvoll insulin sensitivity index; IR, insulin resistance; MPR, matrix pattern recognition; meanRT, reaction time (snap game); MetS, metabolic syndrome; NM, numeric memory; PM, pairs matching test; PAL, paired associated learning test; SDS, symbol digit substitution; TMT, trail-making test (A and B); TRT, tower rearranging; T2D, type 2 diabetes mellitus; WHR, waist-to-hip ratio.

Among the cognitive tests, FI and meanRT showed the higher counts of genomic loci in overlap with IR-related traits, with 111 and 89 significant ρ hits, respectively. Conversely, among the IR-related traits, BMI and WHR displayed the higher numbers of overlapping loci with cognitive tests, with 136 and 94 significant ρ hits. A positive direction of the local bivariate genetic correlations was found in 41% of the significant bivariate tests, with 21 and 19 of the 63 trait pairs carrying only positive or negative local genetic signals. The remaining pairs exhibited a wide dispersion of local effect directions, with negative and positive ρ coexisting in each pair (Supplementary Table S2). In total, 59 of the 221 unique regions were associated with more than one trait pair, henceforth referred to as hotspots (Supplementary Table S3), the major being chr6:32586785-32629239 (between FI and BMI, IFC, ISI, WHR, and between PM and BMI, ISI, and WHR), and chr14:29029225-30831154 (FI-BMI/WHR, NM-BMI/WHR, meanRT-BMI, PM-BMI/HbA1c).

### Fine-mapping of shared genetic association signals

From the 221 candidate regions, mvSuSiE successfully identified a total of 696 high-confidence single-variant credible sets, each one trait-wise significant in at least one cognitive task and one IR-related trait, spanning across 148 genomic regions and involving 54 of the original 110 trait pairs. (Supplementary Table S4). No significant credible sets were found for 55 of the tested regions, while two loci were excluded because fewer than two overlapping SNPs were found between the traits (chr6:28666365-29529755 and chr6:31250557-31320268, both in the MHC region) (44). Figure 2A summarises the number of significant credible sets identified in these hotspots for each trait pair. FI, meanRT, and PM were all characterised by high numbers of shared significant credible sets with BMI and WHR, ranging from 12 to 128 high-confidence causal SNPs. Similarly, MetS shared several significant credible sets with FI (n=19) and meanRT (n=30).

**Figure 2.**
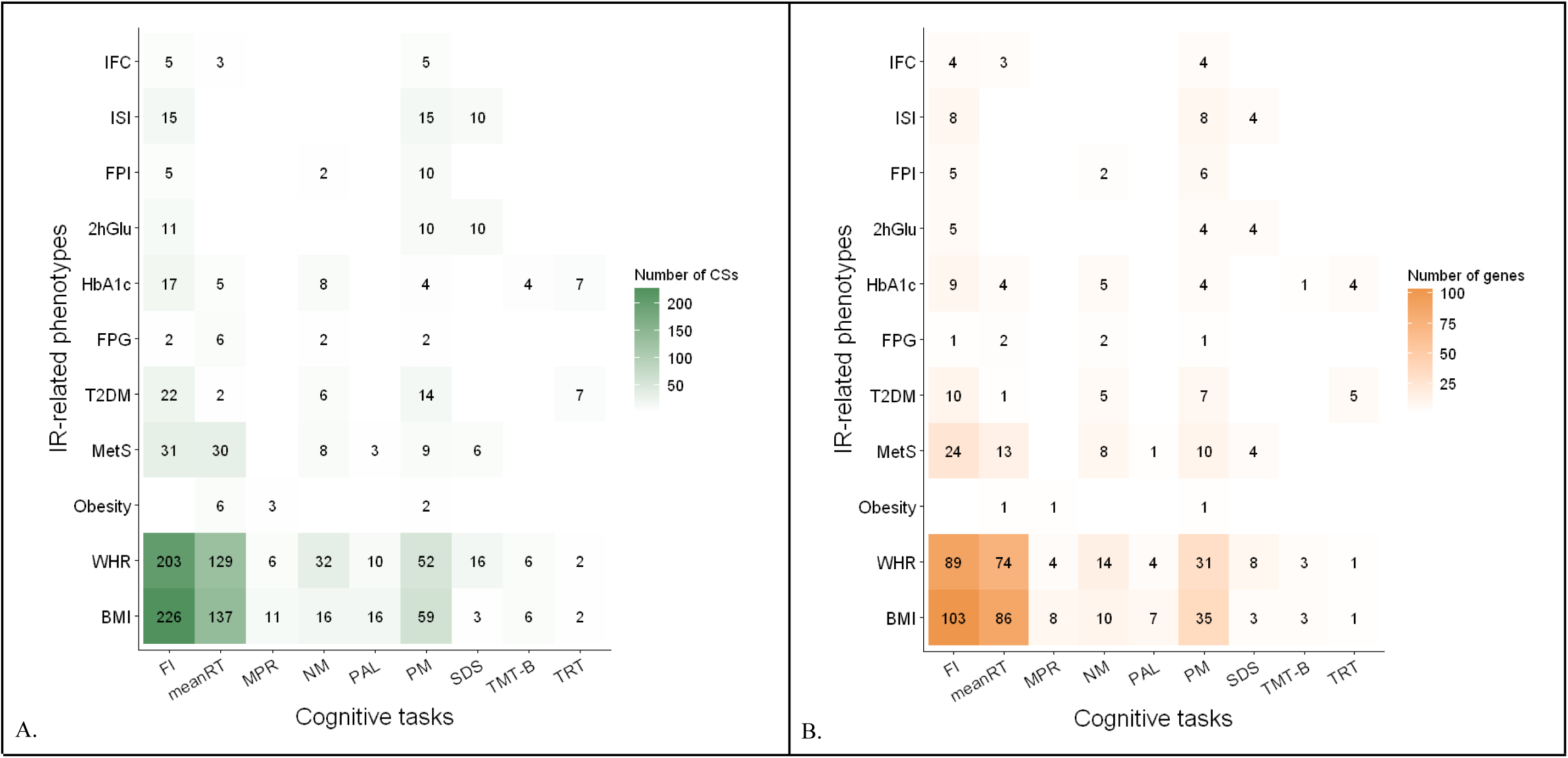
A. Number of credible sets fine-mapped to each pair of traits and **B.** number of relative annotated genes in the credible sets. 2hGlu, 2h glycaemia after an oral glucose test; BMI, body mass index; CS, credible set; FI, fluid intelligence; FPG, fasting plasma glucose; FPI, fasting plasma insulin; HbA1c, glycated haemoglobin; IFC, insulin fold change; ISI, Stumvoll insulin sensitivity index; IR, insulin resistance; MPR, matrix pattern recognition; meanRT, reaction time (snap game); MetS, metabolic syndrome; NM, numeric memory; PM, pairs matching test; PAL, paired associated learning test; SDS, symbol digit substitution; TMT, trail-making test (A and B); TRT, tower rearranging; T2D, type 2 diabetes mellitus; WHR, waist-to-hip ratio.

Fifty-one of the 59 LAVA hotspots were fine mapped to 246 single-variant credible sets (Supplementary Table S5).

### Positional gene mapping

A total of 351 unique genes were positionally mapped from the 696 high-confidence traitwise significant SNPs identified in the mvSuSiE step, of which 458 were mapped through direct overlap with 291 genes, whereas 95 variants mapped to 102 genes within 10 kb window. Most of the identified genes were protein-coding (n=249), followed by long intergenic non-coding RNA (lincRNA, n=42) and antisense (n=26). The number of genes identified per trait pair is summarised in Figure 2B.

A total of 30.9% of the protein-coding genes were shared by more than one trait pair. Pairwise mapped protein coding genes are summarised in Table 2, see Supplementary Tables S6 for complete results.

**Table 2.**
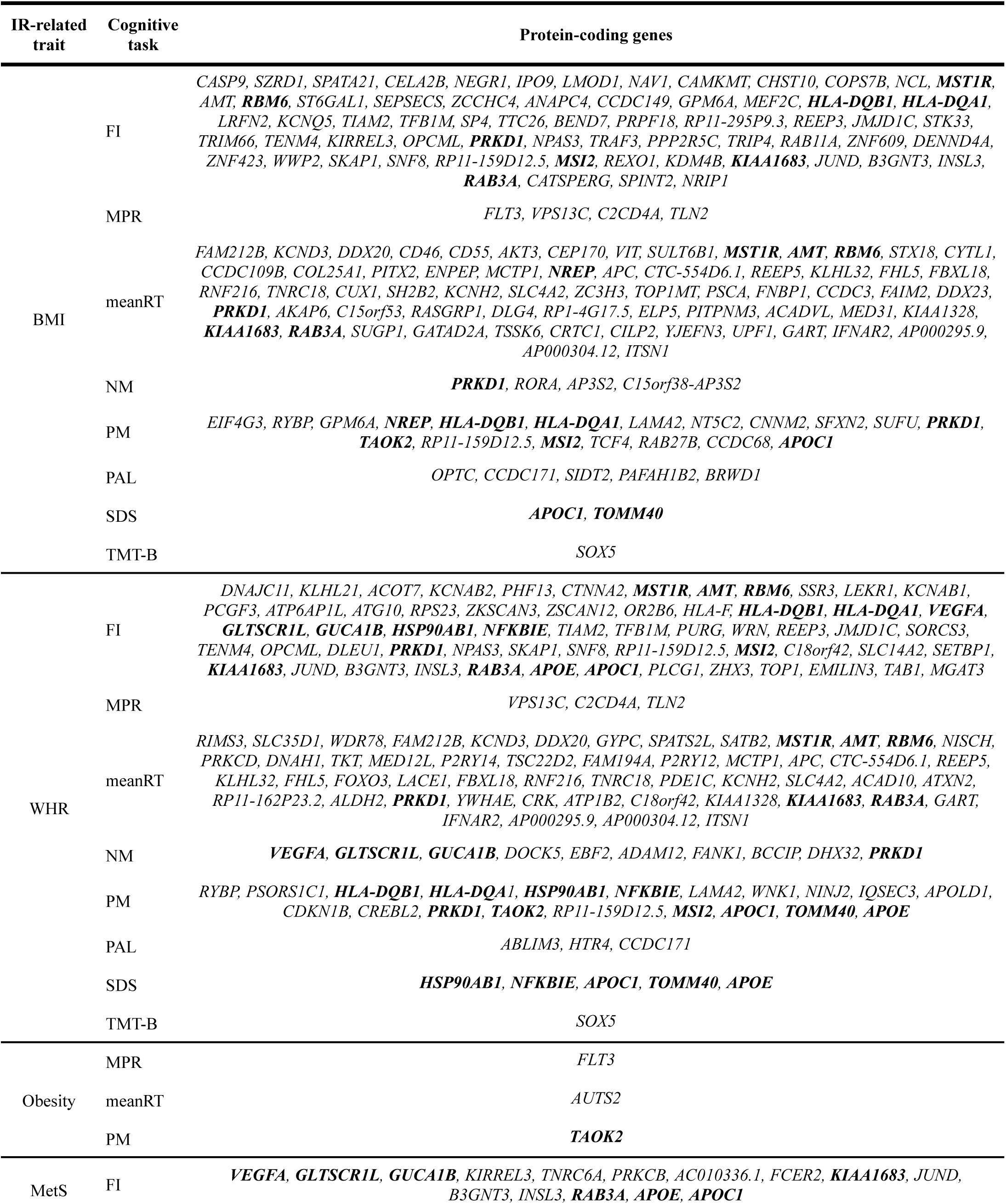
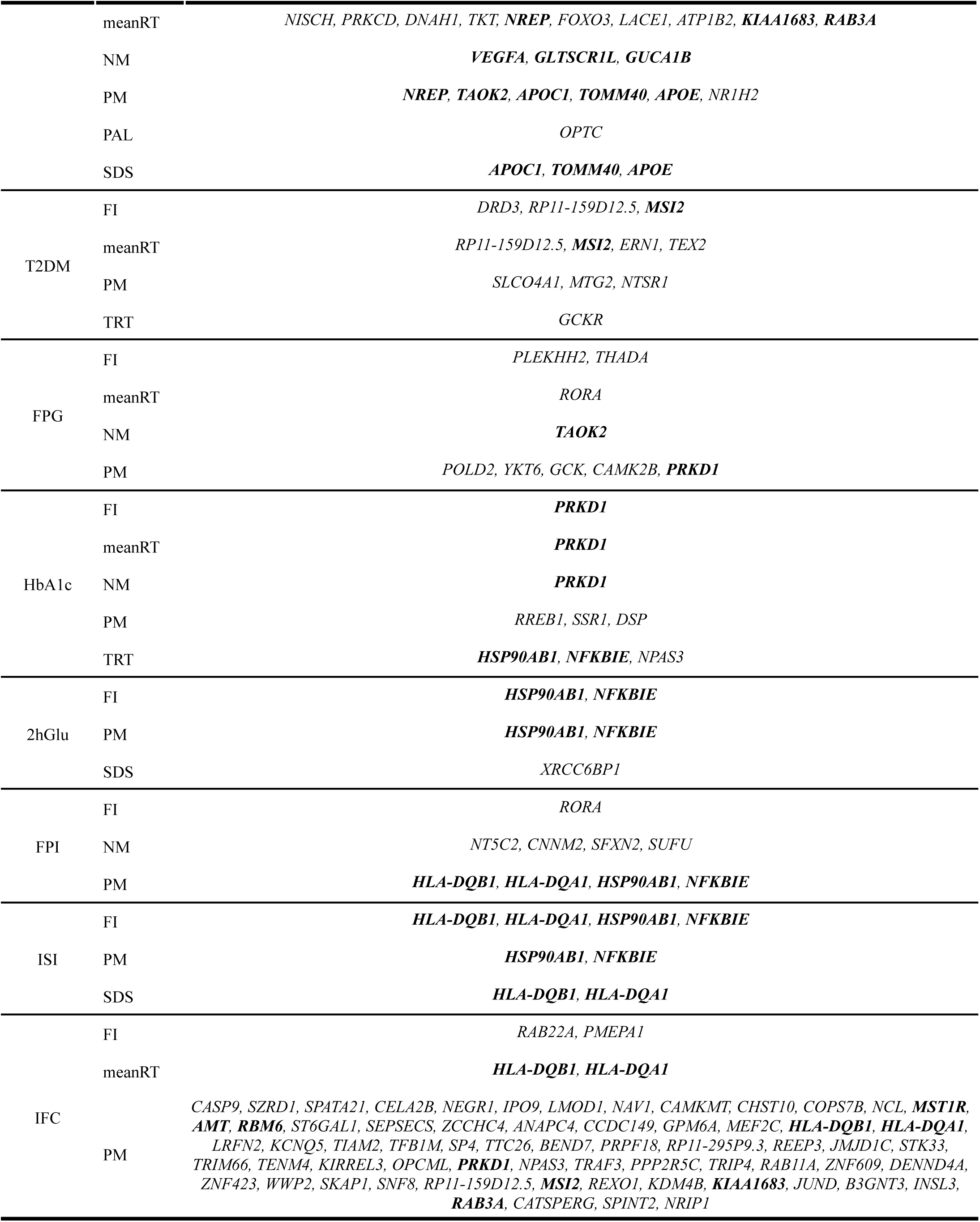
Protein coding genes positionally mapped to each trait pair. 2hGlu, 2h glycaemia after an oral glucose test; BMI, body mass index; FI, fluid intelligence; FPG, fasting plasma glucose; FPI, fasting plasma insulin; HbA1c, glycated haemoglobin; IFC, insulin fold change; ISI, Stumvoll insulin sensitivity index; IR, insulin resistance; MPR, matrix pattern recognition; meanRT, reaction time (snap game); MetS, metabolic syndrome; NM, numeric memory; PM, pairs matching test; PAL, paired associated learning test; SDS, symbol digit substitution; TMT, trail-making test (A and B); TRT, tower rearranging; T2D, type 2 diabetes mellitus; WHR, waist-to-hip ratio. **Bold**: genes recurrent in at least 4 trait pairs.

Comparison of the total list of genes with the NHGRI Catalogue of Published GWAS revealed an enrichment in the genetics of several traits (Supplementary Table S7), like IR-and somatic inflammation-related conditions and indicators. Enrichment was also found for many neuropsychiatric conditions, including Alzheimer’s disease, alcohol consumption and related conditions, attention-deficit/hyperactivity disorder (ADHD), mood and psychotic disorders, as well as for cerebrospinal fluid levels of several proteins and brain imaging measures.

The Genetic Association Database showed an enrichment for variants previously associated with immuno-inflammatory, metabolic, cardiovascular, and oncological diseases, as well as several psychiatric disorders (e.g. ADHD, mood disorders, and schizophrenia) (Supplementary Table S8).

Analysis of of variants found in the Reactome database highlighted an enrichment in SNPs involved in cell cycle regulation, DNA repair, gene expression, immune system regulation, metabolism, signal transduction and neural system-related processes. Enrichment in oncological, inflammatory and infectious processes was also found, consistent with the NHGRI Catalogue findings. See Supplementary Table S9<u>-S10</u> for the full list of associated traits.

### eQTL-gene mapping

A total of 2201 unique genes were mapped through brain eQTLs from the trait pairs that showed the highest number of LAVA-significant loci, *i.e.* BMI-FI, BMI-meanRT, BMI-PM, WHR-FI, WHR-meanRT, WHR-NM, WHR-PM, MetS-FI, and MetS-meanRT (Supplementary Table S11). Overall, 114 genes were mapped for at least four of the above-mentioned traits-pairs and the *PPP4R2* gene recurrently eQTL-mapped across seven trait pairs (BMI-FI/meanRT/PM, WHR-FI/meanRT/PM, and MetS-FI).

### Brain expression and functional characterisation of implicated genes

Positionally mapped genes were enriched among genes differentially expressed in several brain areas, including the BA9 of the frontal cortex, BA24 (corresponding to the anterior cingulate cortex, ACC), hippocampus, amygdala, putamen, caudate nucleus, nucleus accumbens, substantia nigra, cerebellum, and hypothalamus. Other tissues were also enriched, including the heart, ovary, cervix, and gastrointestinal tissues.

Across the pooled set of 2,492 unique positional and eQTL-mapped genes, 971 gene-sets met the prespecified multiple-testing threshold (Supplementary Table S12<u>-S13</u>). Aside from gene-sets related to cognition, brain morphology and metabolism, and IR, significant enrichment was found for gene-sets associated with neuropsychiatric disorders and related phenotypes, immune-inflammatory diseases, and laboratory correlates of cardiometabolic traits. Significant enrichment was also found for pathways related to synaptic and action potential transmission, opioid signaling, neuronal development and plasticity, lipoprotein homeostasis, VEGFR2 signaling, and carcinogenesis. Genes mapped from WHR–PM were enriched for neuronal development and plasticity, and lipidic metabolism, whereas WHR-NM genes were enriched for chemotaxis and vascular regulation. Both FI and PM showed significant enrichment in apolipoprotein metabolism. Additionally, genes mapped from FI-BMI were enriched for macromolecules catabolism, cellular stress response, and neurogenesis. Signal transduction, neurotransmitter regulation, potassium-channels expression, and vesicle-mediated transport were significantly associated with BMI/WHR-meanRT. MetS-FI was significantly enriched for the NR1H3/NR1H2 signaling pathway and lipidic metabolism, together with immune-related gene-sets which were also associated with WHR-PM and WHR-meanRT.

Gene-set enrichment implicated several chromosomic regions, including chr3p13 and chr3p21, chr19p13, chr5q14, and chr6q16, across multiple trait-pair analysis..

Finally, mapped genes showed developmental-stage expression enrichment at 19 post-conceptional weeks (BMI/WHR-PM), 4 years (BMI-FI), 11 years (BMI-meanRT), and 13 years (WHR-PM).

### Drug targetability

Drug-gene interactions were identified for 517 of the 2492 unique positional and eQTL-mapped genes submitted to the DGIdb, with an interaction score ranging from 0.00143 to 104.4. After restricting the interaction score to the fourth quartile (≥1.3) and keeping only the 131 genes in common to at least 4 trait pairs, 45 drug-gene interactions remained between 45 compounds and 22 genes (Supplementary Table S13). Most of the interactions involved 6 genes, i.e. *AMT* (13 drugs), *APOC1* and *GNAI2* (10 drugs), *APEH* and *IMPDH2* (7 drugs), and *COL7A1* (4 drugs) (Table 3). Thirty-two of the identified compounds have reported antineoplastic, anti-inflammatory, or immunomodulating effects, exerted mostly through actions at the sphingosine-1-phosphate receptor (S1PR), the prostaglandin E_2_ receptor (EP2R), the poly-ADP-ribose polymerase 1 (PARP1), and the purinergic receptor P2Y (P2YR).

**Table 3.**
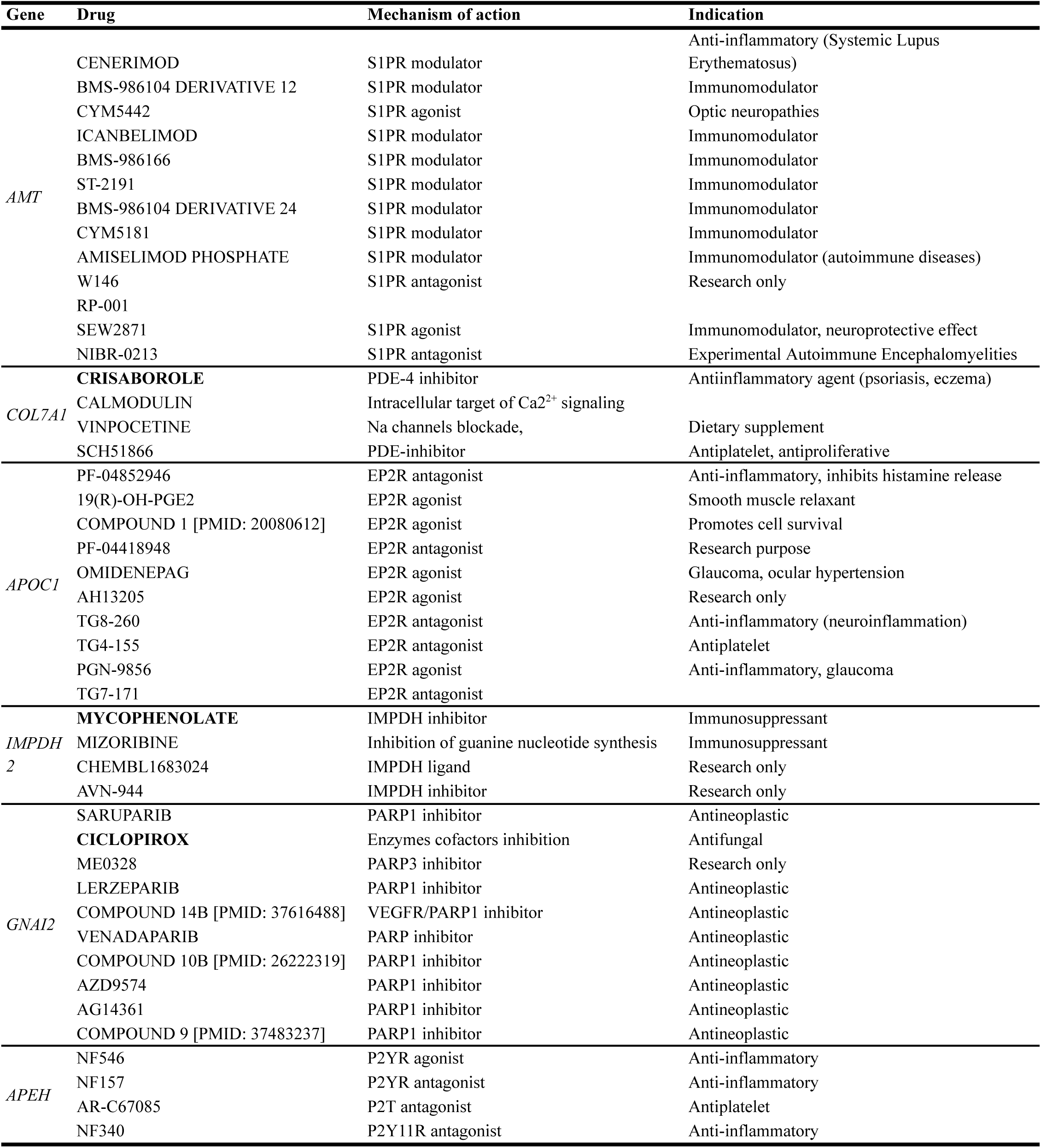
Gene-Drug interaction results from the DGIdb with genes shared by at least 4 trait pairs and with interaction score above the third quartile (1.3). EP2R, prostaglandin E_2_ receptor; IMPDH, inosine 5’-monophosphate dehydrogenase; P2TR, platelet 2T receptor; P2YR, purinergic receptor P2Y; PARP1, poly-ADP-ribose polymerase 1; PARP3, poly(ADP-ribosyl)transferase-3; PDE, phosphodiesterase; S1PR, sphingosine-1-phosphate receptor; VEGFR, vascular endothelial growth factor. **Bold**: drug approved by regulatory agencies.

## Discussion

In this study, we combined genome-wide and local genetic correlation analyses, multivariate fine-mapping, positional and eQTL-based gene mapping, and functional annotation to characterise the shared genetic architecture between insulin resistance (IR)-related traits and cognitive performance. We identified a widespread genetic overlap between cognitive traits and IR-related phenotypes,especially between BMI, WHR and MetS and FI, meanRT, NM, and PM. Mapped genes showed significant enrichment in pathways related to neuronal development and function, immune-inflammatory processes, and lipid and cholesterol metabolism regulation. Implicated genes showed differential expression in the brain at different developmental ages, and had previously been associated with neuropsychiatric and cardiometabolic conditions. These findings support the existence of shared genomic architectures between insulin resistance, cognitive performance, and psychiatric vulnerability, that potentially act across neurodevelopment, and involve immune-related processes and lipid metabolism regulation.

Local genetic correlations showed widespread shared genomic regions, with mixed effect directions in most trait pairs. Together with the evidence of visual declarative short-term memory showing positive global genetic correlation with IR, these results might be read in an evolutionary perspective framing food-seeking behaviours, the ability to forage food, and metabolic resource storage deeply linked to specific cognitive functions (*e.g.* short/long-term memory, processing speed and other executive functions, response inhibition, novel stimuli adaptation) with which may thus share partly overlapping biological determinants (57,58).

Concerning IR-related traits, the predominance in adiposity and body fat distribution traits in our results may have two main explanations. First, from the methodological perspective, body fat distribution traits (WHR and BMI) have bigger GWAS sample sizes and relatively low noise (i.e., definition and measurements are easier), whereas glucose, glycated haemoglobin, and insulin levels are more influenced by acute physiological states, medications, and measurement conditions, and their GWASs are usually corrected for BMI (which might further attenuate the genetic signal). Second, from the biological perspective, adiposity captures a more stable and chronic phenotype, integrating long-term energy balance, lipid storage, inflammatory tone, and endocrine signaling. These features may capture more stable biological exposures relevant to cognitive performance than short-term or context-dependent glycaemic and insulinaemic measures.

Overall, our results suggest that the molecular mechanisms underlying the cognitive-metabolic axis most likely involve lipid and cholesterol metabolism regulation and oxidative stress response, and potentially share a link to the biological basis of common psychiatric disorders. Specific genomic regions appear to be involved, especially the chr19p13 regions. For some trait pairs, functional enrichment appeared to be strongly associated with specific genomic regions, like BMI-meanRT (chr3p21, chr19p13), BMI/MetS-FI (chr19p13), BMI/WHR-PM (chr16p11), and with genes related to lipid metabolism, like *APOE* and *APOC1* (WHR/MetS-FI and BMI/WHR-PM) and the NR1H3/NR1H2 signaling pathway (MetS-FI). The chr16p11 chromosome region has been previously linked to (neurodevelopmental) psychiatric disorders (59–62) and delay discounting (63,64). On the other hand, apolipoproteins are involved in lipid metabolism, participating in the regulation of circulating levels of cholesterol and triglycerides. Apolipoprotein E (ApoE) also has an established critical role in supporting neural function by regulating the efflux of oxidized and harmful metabolites (65–68). The NR1H3/NR1H2 signaling pathway regulates the expression of *APOE* and *APOC* genes in the brain (69) and is involved in lipid metabolism, regulation of cholesterol balance in the brain, neuroinflammation and blood-brain barrier modulation (70,71). *APOE ε4* allele (ApoE4) is the strongest known genetic risk factor for late-onset Alzheimer’s diseases (72,73), but has shown antagonistic pleiotropy features (74) and is linked to improved fitness during foetal development and childhood (73). Although some studies report it to negatively affect intelligence already in childhood and adolescence (75,76), a positive association has also been reported with the *ε4* allele and better cognitive performance (73), especially in visual working memory (77). Finally, ApoE4 has shown to impair insulin receptor trafficking by seizing it in the endosomes, promoting brain insulin resistance (78), and cognitive improvement following intranasal insulin administration in Alzheimer’s disease appears to be mediated by *APOE ε4* allele (79,80). This is consistent with our findings showing a negative global genetic correlation between the IR-related phenotypes and cognition except for the visual declarative short-term memory. Moreover, insulin signaling is known to play a major role in the development of the central nervous system (26), also mediating the effects of early life adverse experiences (31), in line with the evolutionary perspective of the foraging-cognition link. The enrichment found for genes differentially expressed in the brain at different developmental stages suggest that the cognitive-metabolic genetics might also operate during sensitive neurodevelopmental stages, influencing both metabolic regulation and cognitive trajectories.

Taken together, the shared cognitive-metabolic genetic background here characterized might therefore act as one of the central molecular mechanisms underlying neuropsychiatric disorders and the metabolic-psychiatric comorbidity. Early identification of metabolic and cognitive alterations, as well as monitoring these symptoms across clinical trajectories, may support earlier clinical stratification of psychiatric symptoms and disorders, also aided for example by blood markers and cognitive tests measuring specific cognitive functions. Future studies should test whether combined metabolic and cognitive measures improve prediction or clinical stratification beyond established demographic and clinical variables. Moreover, the development of novel therapeutic agents specifically targeting these pathways, if validated through functional *in vitro* analyses, might offer novel treatment options with the ability to address earlier those same symptoms that often represents the highest burden in psychiatric patients (17,19,81,82). Although our drug-gene interaction analyses provide druggability annotations rather than evidence of therapeutic efficacy, they highlight molecular targets that may warrant further experimental investigation. Most of the identified compounds share known anti-inflammatory and immunomodulatory effects. Immune and inflammatory alterations have been reported across several psychiatric disorders and have also been associated with cognitive dysfunction (83–85). For example, APOE is known to modulate the immune system activity, with the *ε4* allele promoting chronic vascular and neuronal inflammation (86,87). Together with other proposed APOE-targeted therapies (88) and immunomodulating approaches (89), the identified molecular pathways, if functionally validated via *in vivo* analyses, may represent future targets for the development of more symptom-tailored and transdiagnostic treatments.

To our knowledge, this is the first study to investigate the shared genetic relationship between multiple IR-related traits and different cognitive functions along the cognitive-metabolic axis through a sequential stepwise framework..

However, this study also comes with some limitations. First, only publicly available data were used, limiting the analyses to a set of traits that do not cover the full range of cognitive functions. To our knowledge, no GWAS summary statistics directly assessing language, complex motor skills, higher order perceptive functions, or social cognition are currently available meeting our inclusion criteria. Second, even though we used the biggest GWASs available, in complex multifactorial conditions and traits even large sample sizes can still be limited, still representing a limiting factor for variant discovery. Third, European ancestries-only GWASs summary statistics were used,restricting our results’ generalizability to other ethnicities. Also, due to the nature of this study, sex-stratified analyses were not possible.

Finally, it is worth noticing that our study does not provide information regarding causality. Experimental and preclinical follow-up studies are needed to further probe the molecular mechanisms identified, whilst longitudinal approaches should be considered to assess causality and phenotype changes across the years. Clinical implications of our results should be further explored, too, especially for neuropsychiatric disorders.

In conclusion, heterogeneous shared genetic architecture exists between IR and cognitive traits, especially involving adiposity-related phenotypes and measures of general intelligence, processing speed and short-term declarative memory. Neuronal, immune-inflammatory, and lipid-metabolism processes were implicated, with drug-gene analyses prioritising molecular targets for future experimental investigations. Our findings provide a detailed genetic characterization of the cognitive-metabolic axis, paving the way for future mechanistic studies.

## Data Availability

Main data produced in the present work are contained in the manuscript, in the Supplementary Files. Additionally, raw data are available upon reasonable request to the authors.
LAVA and mvSuSiE codes used in this study are available on GitHub: https://github.com/AlphMars/LAVA-parallelisation, https://github.com/AlphMars/mvSuSiE-parallelisation.

https://github.com/AlphMars/mvSuSiE-parallelisation

## Acknowledgements

Janita Bralten was supported by a personal grant from the Netherlands Organization for Scientific Research (NWO) Innovation Program (Vidi grant No. 09150172410043). Research reported in this publication was supported by the National Institute Of Mental Health of the National Institutes of Health under Award Number R01MH124851. The content is solely the responsibility of the authors and does not necessarily represent the official views of the National Institutes of Health. This work used the Dutch national e-infrastructure with the support of the SURF Cooperative using grant no. EINF-13722. We also thank the researchers of the consortia that provided the GWAS summary statistics used in our analyses and the participants of the cohorts to which they refer.

## Code and data availability

LAVA and mvSuSiE codes used in this study are available on GitHub: https://github.com/AlphMars/LAVA-parallelisation,

https://github.com/AlphMars/mvSuSiE-parallelisation.

GWAS summary statistics of the cognitive tasks were obtained upon direct request to the authors, for IR-related traits source websites are reported in Supplementary Table S1.

## Declaration of Generative AI and AI-assisted technologies in the writing process

The authors used OpenAI’s ChatGPT 5.6 for language editing and improvement of sentence formulation. No scientific content, interpretation, literature selection, or conclusions were generated by the tool. The authors critically reviewed and revised all artificial intelligence-assisted text and take full responsibility for the accuracy, interpretation, and final wording of the manuscript.

## References

1. Lee SH, Park SY, Choi CS. Insulin Resistance: From Mechanisms to Therapeutic Strategies. Diabetes Metab J. 2022 Jan;46(1):15–37. doi:10.4093/dmj.2021.0280 PubMed PMID: 34965646; PubMed Central PMCID: PMC8831809.

2. Fanelli G, Mota NR, Salas-Salvadó J, Bulló M, Fernandez-Aranda F, Camacho-Barcia L, et al. The link between cognition and somatic conditions related to insulin resistance in the UK Biobank study cohort: a systematic review. Neuroscience & Biobehavioral Reviews. 2022 Dec 1;143:104927. doi:10.1016/j.neubiorev.2022.104927

3. Garfield V, Farmaki AE, Eastwood SV, Mathur R, Rentsch CT, Bhaskaran K, et al. HbA1c and brain health across the entire glycaemic spectrum. Diabetes, Obesity and Metabolism. 2021 May;23(5):1140–9. doi:10.1111/dom.14321

4. Gruber JR, Ruf A, Süß ED, Tariverdian S, Ahrens KF, Schiweck C, et al. Impact of blood glucose on cognitive function in insulin resistance: novel insights from ambulatory assessment. Nutr Diabetes. 2024 Sep 11;14(1):74. doi:10.1038/s41387-024-00331-0

5. Lyall DM, Celis-Morales CA, Anderson J, Gill JMR, Mackay DF, McIntosh AM, et al. Associations between single and multiple cardiometabolic diseases and cognitive abilities in 474 129 UK Biobank participants. Eur Heart J. 2017 Feb 21;38(8):577–83. doi:10.1093/eurheartj/ehw528

6. Menon AJ, Selva M, Sandhya G, Singh S, Abhishek ML, Stezin A, et al. Understanding the link between insulin resistance and cognition: a cross-sectional study conducted in an urban, South Indian cohort. Acta Diabetol. 2025 Sep 1;62(9):1507–14. doi:10.1007/s00592-025-02483-6

7. Newby D, Garfield V. Understanding the inter-relationships of type 2 diabetes and hypertension with brain and cognitive health: A UK Biobank study. Diabetes, Obesity and Metabolism. 2022 May;24(5):938–47. doi:10.1111/dom.14658

8. Li C, Gheorghe DA, Gallacher JE, Bauermeister S. Psychiatric comorbid disorders of cognition: a machine learning approach using 1175 UK Biobank participants. Evid Based Mental Health. 2020 Oct 28;23(4). doi:10.1136/ebmental-2020-300147 PubMed PMID: 10.1136/ebmental-2020-300147.

9. Ferguson AC, Tank R, Lyall LM, Ward J, Welsh P, Celis-Morales C, et al. Association of SBP and BMI with cognitive and structural brain phenotypes in UK Biobank. Journal of Hypertension. 2020 Dec;38(12):2482. doi:10.1097/HJH.0000000000002579

10. Klinedinst BS, Pappas C, Le S, Yu S, Wang Q, Wang L, et al. Aging-related changes in fluid intelligence, muscle and adipose mass, and sex-specific immunologic mediation: A longitudinal UK Biobank study. Brain, Behavior, and Immunity. 2019 Nov 1;82:396–405. doi:10.1016/j.bbi.2019.09.008

11. Morys F, Dadar M, Dagher A. Association Between Midlife Obesity and Its Metabolic Consequences, Cerebrovascular Disease, and Cognitive Decline. J Clin Endocrinol Metab. 2021 Oct 1;106(10):e4260–74. doi:10.1210/clinem/dgab135

12. Olivo G, Gour S, Schiöth HB. Low neuroticism and cognitive performance are differently associated to overweight and obesity: A cross-sectional and longitudinal UK Biobank study. Psychoneuroendocrinology. 2019 Mar 1;101:167–74. doi:10.1016/j.psyneuen.2018.11.014

13. Brown AE, Walker M. Genetics of Insulin Resistance and the Metabolic Syndrome. Curr Cardiol Rep. 2016;18:75. doi:10.1007/s11886-016-0755-4 PubMed PMID: 27312935; PubMed Central PMCID: PMC4911377.

14. Oliveri A, Rebernick RJ, Kuppa A, Pant A, Chen Y, Du X, et al. Comprehensive genetic study of the insulin resistance marker TG:HDL-C in the UK Biobank. Nat Genet. 2024 Feb;56(2):212–21. doi:10.1038/s41588-023-01625-2

15. Spoto G, Di Rosa G, Nicotera AG. The Impact of Genetics on Cognition: Insights into Cognitive Disorders and Single Nucleotide Polymorphisms. J Pers Med. 2024 Jan 30;14(2):156. doi:10.3390/jpm14020156 PubMed PMID: 38392589; PubMed Central PMCID: PMC10889941.

16. Wootton O, Shadrin AA, Bjella T, Smeland OB, van der Meer D, Frei O, et al. Genomic insights into the shared and distinct genetic architecture of cognitive function and schizophrenia. Sci Rep. 2024 Jul 4;14(1):15356. doi:10.1038/s41598-024-66085-y

17. Wang Y, Meng W, Liu Z, An Q, Hu X. Cognitive impairment in psychiatric diseases: Biomarkers of diagnosis, treatment, and prevention. Front Cell Neurosci. 2022 Nov 2;16. doi:10.3389/fncel.2022.1046692

18. Wimberley T, Horsdal HT, Brikell I, Laursen TM, Astrup A, Fanelli G, et al. Temporally ordered associations between type 2 diabetes and brain disorders – a Danish register-based cohort study. BMC Psychiatry. 2022 Aug 26;22(1):573. doi:10.1186/s12888-022-04163-z

19. Penadés R, Forte MF, Mezquida G, Andrés C, Catalán R, Segura B. Treating Cognition in Schizophrenia: A Whole Lifespan Perspective. Healthcare (Basel). 2024 Nov 4;12(21):2196. doi:10.3390/healthcare12212196 PubMed PMID: 39517406; PubMed Central PMCID: PMC11545462.

20. Schandorff JM, Damgaard V, Little B, Kjærstad HL, Zarp J, Bjertrup AJ, et al. Cognitive hierarchy in mood disorders and relations to daily functioning. Journal of Affective Disorders. 2025 Apr;375:239–48. doi:10.1016/j.jad.2025.01.143

21. Tsapekos D, Kalfas M, Strawbridge R, Swidzinski S, Burdick KE, Young AH. Estimating Cognitive Impairment in Bipolar Disorder: Should We Account for Premorbid IQ ? Acta Psychiatr Scand. 2025 Jun 18;acps.70000. doi:10.1111/acps.70000

22. Lyoo IK, Yoon S, Renshaw PF, Hwang J, Bae S, Musen G, et al. Network-Level Structural Abnormalities of Cerebral Cortex in Type 1 Diabetes Mellitus. Zuo XN, editor. PLoS ONE. 2013 Aug 23;8(8):e71304. doi:10.1371/journal.pone.0071304

23. Nevo-Shenker M, Shalitin S. The Impact of Hypo-and Hyperglycemia on Cognition and Brain Development in Young Children with Type 1 Diabetes. Horm Res Paediatr. 2021 Jul 9;94(3–4):115–23. doi:10.1159/000517352

24. Awad N, Gagnon M, Messier C. The Relationship between Impaired Glucose Tolerance, Type 2 Diabetes, and Cognitive Function. Journal of Clinical and Experimental Neuropsychology. 2004 Nov 1;26(8):1044–80. doi:10.1080/13803390490514875 PubMed PMID: 15590460.

25. Messier C, Awad-Shimoon N, Gagnon M, Desrochers A, Tsiakas M. Glucose regulation is associated with cognitive performance in young nondiabetic adults. Behavioural Brain Research. 2011 Sep 12;222(1):81–8. doi:10.1016/j.bbr.2011.03.023

26. Réthelyi JM, Vincze K, Schall D, Glennon J, Berkel S. The role of insulin/IGF1 signalling in neurodevelopmental and neuropsychiatric disorders – Evidence from human neuronal cell models. Neuroscience & Biobehavioral Reviews. 2023 Oct 1;153:105330. doi:10.1016/j.neubiorev.2023.105330

27. Sullivan M, Fernandez-Aranda F, Camacho-Barcia L, Harkin A, Macrì S, Mora-Maltas B, et al. Insulin and disorders of behavioural flexibility. Neuroscience & Biobehavioral Reviews. 2023 Jul 1;150:105169. doi:10.1016/j.neubiorev.2023.105169

28. Gruber J, Hanssen R, Qubad M, Bouzouina A, Schack V, Sochor H, et al. Impact of insulin and insulin resistance on brain dopamine signalling and reward processing – An underexplored mechanism in the pathophysiology of depression? Neuroscience & Biobehavioral Reviews. 2023 Jun 1;149:105179. doi:10.1016/j.neubiorev.2023.105179

29. Fanelli G, Franke B, Fabbri C, Werme J, Erdogan I, De Witte W, et al. Local patterns of genetic sharing between neuropsychiatric and insulin resistance-related conditions. Transl Psychiatry. 2025 Apr 12;15(1):1–10. doi:10.1038/s41398-025-03349-9

30. Possidente C, Fanelli G, Serretti A, Fabbri C. Clinical insights into the cross-link between mood disorders and type 2 diabetes: A review of longitudinal studies and Mendelian randomisation analyses. Neuroscience & Biobehavioral Reviews. 2023 Sep 1;152:105298. doi:10.1016/j.neubiorev.2023.105298

31. Alberry B, Silveira PP. Brain insulin signaling as a potential mediator of early life adversity effects on physical and mental health. Neuroscience & Biobehavioral Reviews. 2023 Oct 1;153:105350. doi:10.1016/j.neubiorev.2023.105350

32. Wu D, Li Y, Chen L, Klein M, Franke B, Chen J, et al. Maternal gestational weight gain and offspring’s neurodevelopmental outcomes: A systematic review and meta-analysis. Neuroscience & Biobehavioral Reviews. 2023 Oct 1;153:105360. doi:10.1016/j.neubiorev.2023.105360

33. Nolan E, Sawyer A, Archer DB, Dumitrescu L, Kukull WW, Biber S, et al. Genetic associations between insulin resistance and memory performance/decline: A NACC cohort study. Alzheimers Dement. 2025 Dec 23;21(Suppl 6):e106817. doi:10.1002/alz70860_106817 PubMed PMID: 41434115; PubMed Central PMCID: PMC12725150.

34. Sarnowski C, Zhang Y, Ammous F, Shade LMP, DiCorpo D, Jian X, et al. Association of genetic scores related to insulin resistance with neurological outcomes in ancestrally diverse cohorts from the Trans-Omics for Precision Medicine (TOPMed) program. Commun Biol. 2025 Sep 24;8(1):1352. doi:10.1038/s42003-025-08674-9

35. Su F, Shu H, Ye Q, Wang Z, Xie C, Yuan B, et al. Brain insulin resistance deteriorates cognition by altering the topological features of brain networks. Neuroimage Clin. 2016 Dec 12;13:280–7. doi:10.1016/j.nicl.2016.12.009 PubMed PMID: 28050343; PubMed Central PMCID: PMC5192246.

36. Fawns-Ritchie C, Deary IJ. Reliability and validity of the UK Biobank cognitive tests. PLOS ONE. 2020 Apr 20;15(4):e0231627. doi:10.1371/journal.pone.0231627

37. Rudvik A, Månsson M. Evaluation of surrogate measures of insulin sensitivity - correlation with gold standard is not enough. BMC Med Res Methodol. 2018 Jun 26;18:64. doi:10.1186/s12874-018-0521-y PubMed PMID: 29940866; PubMed Central PMCID: PMC6019831.

38. Grotzinger AD, Rhemtulla M, de Vlaming R, Ritchie SJ, Mallard TT, Hill WD, et al. Genomic structural equation modelling provides insights into the multivariate genetic architecture of complex traits. Nat Hum Behav. 2019 May;3(5):513–25. doi:10.1038/s41562-019-0566-x

39. Bulik-Sullivan BK, Loh PR, Finucane HK, Ripke S, Yang J, Patterson N, et al. LD Score regression distinguishes confounding from polygenicity in genome-wide association studies. Nat Genet. 2015 Mar;47(3):291–5. doi:10.1038/ng.3211

40. The 1000 Genomes Project Consortium, Corresponding authors, Auton A, Abecasis GR, Steering committee, Altshuler DM, et al. A global reference for human genetic variation. Nature. 2015 Oct 1;526(7571):68–74. doi:10.1038/nature15393

41. Werme J, van der Sluis S, Posthuma D, de Leeuw CA. An integrated framework for local genetic correlation analysis. Nat Genet. 2022 Mar;54(3):274–82. doi:10.1038/s41588-022-01017-y

42. Cadeleeuw. cadeleeuw/lava-partitioning: [Internet]. Zenodo; 2021 [cited 2025 Aug 22]. Available from: https://zenodo.org/record/5583779 doi:10.5281/ZENODO.5583779

43. Reynolds RH, Wagen AZ, Lona-Durazo F, Scholz SW, Shoai M, Hardy J, et al. Local genetic correlations exist among neurodegenerative and neuropsychiatric diseases. npj Parkinsons Dis. 2023 Apr 28;9(1):70. doi:10.1038/s41531-023-00504-1

44. Zou Y, Carbonetto P, Xie D, Wang G, Stephens M. Fast and flexible joint fine-mapping of multiple traits via the Sum of Single Effects model [Internet]. bioRxiv; 2024 [cited 2025 Aug 24]. p. 2023.04.14.536893. Available from: https://www.biorxiv.org/content/10.1101/2023.04.14.536893v5doi:10.1101/2023.04.14.536893

45. Wang G, Sarkar A, Carbonetto P, Stephens M. A Simple New Approach to Variable Selection in Regression, with Application to Genetic Fine Mapping. J R Stat Soc Ser B Stat Methodol. 2020 Dec 1;82(5):1273–300. doi:10.1111/rssb.12388

46. Urbut SM, Wang G, Carbonetto P, Stephens M. Flexible statistical methods for estimating and testing effects in genomic studies with multiple conditions. Nat Genet. 2019 Jan;51(1):187–95. doi:10.1038/s41588-018-0268-8 PubMed PMID: 30478440; PubMed Central PMCID: PMC6309609.

47. Dayem Ullah AZ, Lemoine NR, Chelala C. A practical guide for the functional annotation of genetic variations using SNPnexus. Brief Bioinform. 2013 Jul 1;14(4):437–47. doi:10.1093/bib/bbt004

48. Oscanoa J, Sivapalan L, Gadaleta E, Dayem Ullah AZ, Lemoine NR, Chelala C. SNPnexus: a web server for functional annotation of human genome sequence variation (2020 update). Nucleic Acids Res. 2020 Jul 2;48(W1):W185–92. doi:10.1093/nar/gkaa420

49. Xu T, Jin P, Qin ZS. Regulatory annotation of genomic intervals based on tissue-specific expression QTLs. Bioinformatics. 2020 Feb 1;36(3):690–7. doi:10.1093/bioinformatics/btz669

50. Watanabe K, Taskesen E, van Bochoven A, Posthuma D. Functional mapping and annotation of genetic associations with FUMA. Nat Commun. 2017 Nov 28;8(1):1826. doi:10.1038/s41467-017-01261-5

51. Cannon M, Stevenson J, Stahl K, Basu R, Coffman A, Kiwala S, et al. DGIdb 5.0: rebuilding the drug-gene interaction database for precision medicine and drug discovery platforms. Nucleic Acids Res. 2024 Jan 5;52(D1):D1227–35. doi:10.1093/nar/gkad1040 PubMed PMID: 37953380; PubMed Central PMCID: PMC10767982.

52. Chen J, Spracklen CN, Marenne G, Varshney A, Corbin LJ, Luan J, et al. The trans-ancestral genomic architecture of glycemic traits. Nat Genet. 2021 Jun;53(6):840–60. doi:10.1038/s41588-021-00852-9

53. Lind L. Genome-Wide Association Study of the Metabolic Syndrome in UK Biobank. Metab Syndr Relat Disord. 2019 Dec;17(10):505–11. doi:10.1089/met.2019.0070 PubMed PMID: 31589552.

54. Mahajan A, Spracklen CN, Zhang W, Ng MCY, Petty LE, Kitajima H, et al. Multi-ancestry genetic study of type 2 diabetes highlights the power of diverse populations for discovery and translation. Nat Genet. 2022 May;54(5):560–72. doi:10.1038/s41588-022-01058-3

55. Pulit SL, Stoneman C, Morris AP, Wood AR, Glastonbury CA, Tyrrell J, et al. Meta-analysis of genome-wide association studies for body fat distribution in 694 649 individuals of European ancestry. Hum Mol Genet. 2019 Jan 1;28(1):166–74. doi:10.1093/hmg/ddy327

56. Williamson A, Norris DM, Yin X, Broadaway KA, Moxley AH, Vadlamudi S, et al. Genome-wide association study and functional characterization identifies candidate genes for insulin-stimulated glucose uptake. Nat Genet. 2023 Jun;55(6):973–83. doi:10.1038/s41588-023-01408-9 PubMed PMID: 37291194; PubMed Central PMCID: PMC7614755.

57. Hayden BY, Walton ME. Neuroscience of foraging. Front Neurosci. 2014 Apr 21;8. doi:10.3389/fnins.2014.00081

58. Wu T, Xu S. Understanding the contemporary high obesity rate from an evolutionary genetic perspective. Hereditas. 2023 Feb 7;160(1):5. doi:10.1186/s41065-023-00268-x

59. Gregersen NO, Buttenschøn HN, Hedemand A, Nielsen MN, Dahl HA, Kristensen AS, et al. Association between genes on chromosome 19p13.2 and panic disorder. Psychiatric Genetics. 2016 Dec;26(6):287. doi:10.1097/YPG.0000000000000147

60. Hanson E, Bernier R, Porche K, Jackson FI, Goin-Kochel RP, Snyder LG, et al. The Cognitive and Behavioral Phenotype of the 16p11.2 Deletion in a Clinically Ascertained Population. Biological Psychiatry. 2015 May 1;Autism Genotypes and Phenotypes77(9):785–93. doi:10.1016/j.biopsych.2014.04.021

61. Hayashi Y, Kushima I, Aleksic B, Senaha T, Ozaki N. Variable psychiatric manifestations in patients with 16p11.2 duplication: a case series of 4 patients. Psychiatry and Clinical Neurosciences. 2022;76(3):86–8. doi:10.1111/pcn.13324

62. Taylor CM, Smith R, Lehman C, Mitchel MW, Singer K, Weaver WC, et al. 16p11.2 Recurrent Deletion. In: Adam MP, Bick S, Mirzaa GM, Pagon RA, Wallace SE, Amemiya A, editors. GeneReviews® [Internet]. Seattle (WA): University of Washington, Seattle; 1993 [cited 2025 Dec 6]. Available from: http://www.ncbi.nlm.nih.gov/books/NBK11167/ PubMed PMID: 20301775.

63. Pirim D, Radwan ZH, Wang X, Niemsiri V, Hokanson JE, Hamman RF, et al. Apolipoprotein E-C1-C4-C2 gene cluster region and inter-individual variation in plasma lipoprotein levels: a comprehensive genetic association study in two ethnic groups. PLOS ONE. 2019 Mar 26;14(3):e0214060. doi:10.1371/journal.pone.0214060

64. Thorpe HHA, Cupertino RB, Pakala SR, Fontanillas P, Jennings MV, Yang J, et al. Genome-wide association study of delay discounting identifies 11 loci and reveals transdiagnostic associations across mental and physical health. Mol Psychiatry. 2025 Nov 25;1–13. doi:10.1038/s41380-025-03356-8

65. Abondio P, Sazzini M, Garagnani P, Boattini A, Monti D, Franceschi C, et al. The Genetic Variability of APOE in Different Human Populations and Its Implications for Longevity. Genes (Basel). 2019 Mar 15;10(3):222. doi:10.3390/genes10030222 PubMed PMID: 30884759; PubMed Central PMCID: PMC6471373.

66. APOE Gene - GeneCards [Internet]. [cited 2025 Dec 6]. Available from: https://www.genecards.org/cgi-bin/carddisp.pl?gene=APOE&keywords=APOE

67. Lyall DM, Celis-Morales C, Lyall LM, Graham C, Graham N, Mackay DF, et al. Assessing for interaction between APOE ε4, sex, and lifestyle on cognitive abilities. Neurology. 2019 Jun 4;92(23):e2691–8. doi:10.1212/WNL.0000000000007551

68. Ralhan I, Do AD, Bae JY, Feringa FM, Cai W, Chang J, et al. Protective ApoE variants support neuronal function by effluxing oxidized phospholipids. Neuron. 2025 Dec 2;0(0). doi:10.1016/j.neuron.2025.10.040 PubMed PMID: 41338186.

69. Fessler MB. Liver X Receptor: Crosstalk Node for the Signaling of Lipid Metabolism, Carbohydrate Metabolism, and Innate Immunity. Curr Signal Transduct Ther. 2008 May 1;3(2):75–81. doi:10.2174/157436208784223170 PubMed PMID: 24563635; PubMed Central PMCID: PMC3931522.

70. Cummins C, Repa J, Matthews L, Shamovsky V. NR1H2 and NR1H3-mediated signaling [Internet]. 2019 [cited 2025 Dec 6]. Available from: http://reactome.org/content/detail/R-HSA-9024446.1 doi:10.3180/R-HSA-9024446.1

71. Vacondio D, Nogueira Pinto H, Coenen L, Mulder IA, Fontijn R, van Het Hof B, et al. Liver X receptor alpha ensures blood-brain barrier function by suppressing SNAI2. Cell Death Dis. 2023 Nov 28;14(11):781. doi:10.1038/s41419-023-06316-8 PubMed PMID: 38016947; PubMed Central PMCID: PMC10684660.

72. Liu CC, Kanekiyo T, Xu H, Bu G. Apolipoprotein E and Alzheimer disease: risk, mechanisms and therapy. Nat Rev Neurol. 2013 Feb;9(2):106–18. doi:10.1038/nrneurol.2012.263

73. Smith CJ, Ashford JW, Perfetti TA. Putative Survival Advantages in Young Apolipoprotein ɛ4 Carriers are Associated with Increased Neural Stress. J Alzheimers Dis. 2019;68(3):885–923. doi:10.3233/JAD-181089 PubMed PMID: 30814349; PubMed Central PMCID: PMC6484250.

74. Tuminello ER, Han SD. The apolipoprotein e antagonistic pleiotropy hypothesis: review and recommendations. Int J Alzheimers Dis. 2011 Feb 24;2011:726197. doi:10.4061/2011/726197 PubMed PMID: 21423560; PubMed Central PMCID: PMC3056453.

75. Bloss CS, Delis DC, Salmon DP, Bondi MW. APOE genotype is associated with left-handedness and visuospatial skills in children. Neurobiology of Aging. 2010 May 1;31(5):787–95. doi:10.1016/j.neurobiolaging.2008.05.021

76. Reynolds CA, Smolen A, Corley RP, Munoz E, Friedman NP, Rhee SH, et al. APOE effects on cognition from childhood to adolescence. Neurobiology of Aging. 2019 Dec 1;84:239.e1–239.e8. doi:10.1016/j.neurobiolaging.2019.04.011

77. Lu K, Nicholas JM, Pertzov Y, Grogan J, Husain M, Pavisic IM, et al. Dissociable effects of APOE ε4 and β-amyloid pathology on visual working memory. Nat Aging. 2021 Nov;1(11):1002–9. doi:10.1038/s43587-021-00117-4

78. Zhao N, Liu CC, Ingelgom AJV, Martens YA, Linares C, Knight JA, et al. Apolipoprotein E4 Impairs Neuronal Insulin Signaling by Trapping Insulin Receptor in the Endosomes. Neuron. 2017 Sep 27;96(1):115–129.e5. doi:10.1016/j.neuron.2017.09.003 PubMed PMID: 28957663.

79. Hallschmid M. Intranasal insulin. Journal of Neuroendocrinology. 2021;33(4):e12934. doi:10.1111/jne.12934

80. Tabassum A, Badulescu S, Singh E, Asoro R, McIntyre RS, Teopiz KM, et al. Central effects of acute intranasal insulin on neuroimaging, cognitive, and behavioural outcomes: A systematic review. Neuroscience & Biobehavioral Reviews. 2024 Dec 1;167:105907. doi:10.1016/j.neubiorev.2024.105907

81. Huang CLC. Residual Cognitive Deficit in Adults with Depression who Recovered after 6-month Treatment: Stable versus State-Dependent Markers. J Clin Med Res. 2009 Oct;1(4):202–6. doi:10.4021/jocmr2009.10.1266 PubMed PMID: 22461869; PubMed Central PMCID: PMC3299181.

82. Sumiyoshi T, Hoshino T, Mishiro I, Hammer-Helmich L, Ge H, Moriguchi Y, et al. Prediction of residual cognitive disturbances by early response of depressive symptoms to antidepressant treatments in patients with major depressive disorder. J Affect Disord. 2022 Jan 1;296:95–102. doi:10.1016/j.jad.2021.09.025 PubMed PMID: 34597893.

83. Ferat-Osorio E, Maldonado-García JL, Pavón L. How inflammation influences psychiatric disease. World J Psychiatry. 2024 Mar 19;14(3):342–9. doi:10.5498/wjp.v14.i3.342 PubMed PMID: 38617981; PubMed Central PMCID: PMC11008389.

84. Tan S, Chen W, Kong G, Wei L, Xie Y. Peripheral inflammation and neurocognitive impairment: correlations, underlying mechanisms, and therapeutic implications. Front Aging Neurosci. 2023 Nov 29;15. doi:10.3389/fnagi.2023.1305790

85. Zhao F, Li B, Yang W, Ge T, Cui R. Brain-immune interaction mechanisms: Implications for cognitive dysfunction in psychiatric disorders. Cell Prolif. 2022 Oct;55(10):e13295. doi:10.1111/cpr.13295 PubMed PMID: 35860850; PubMed Central PMCID: PMC9528770.

86. Yin C, Ackermann S, Ma Z, Mohanta SK, Zhang C, Li Y, et al. ApoE attenuates unresolvable inflammation by complex formation with activated C1q. Nat Med. 2019 Mar;25(3):496–506. doi:10.1038/s41591-018-0336-8

87. Parhizkar S, Holtzman DM. APOE mediated neuroinflammation and neurodegeneration in Alzheimer’s disease. Seminars in Immunology. 2022 Jan 1;59:101594. doi:10.1016/j.smim.2022.101594

88. Yassine HN, Hugo C, O’Donovan B, Stephens IO, Johnson LA, Cole G, et al. APOE-Targeted Therapeutics for Alzheimer’s Disease. J Neurosci. 2025 Nov 12;45(46):e1388252025. doi:10.1523/JNEUROSCI.1388-25.2025 PubMed PMID: 41224653; PubMed Central PMCID: PMC12614056.

89. Fox NC, Belder C, Ballard C, Kales HC, Mummery C, Caramelli P, et al. Treatment for Alzheimer’s disease. The Lancet. 2025 Sep 27;406(10510):1408–23. doi:10.1016/S0140-6736(25)01329-7 PubMed PMID: 40997839.

